# Association between postoperative fine particulate matter exposure and right ventricle-pulmonary artery conduit patency

**DOI:** 10.1101/2023.03.21.23287552

**Authors:** Chao Yue, Jin Li, Jiaqi Zhang, Qiang Wang, Xu Wang

**Affiliations:** Department of Pediatric Cardiac Center, National Center for Cardiovascular Diseases, Fuwai Hospital, Chinese Academy of Medical Sciences and Peking Union Medical College, No. 167 Beilishi Street, Xicheng District, Beijing 100037, China; State key joint laboratory of Environment Simulation and Pollution Control, School of Environment, Tsinghua University, No.30 Shuangqing Street, Haidian District, Beijing 100084, China; Department of Echocardiography, National Center for Cardiovascular Diseases, Fuwai Hospital, Chinese Academy of Medical Sciences and Peking Union Medical College, No. 167 Beilishi Street, Xicheng District, Beijing 100037, China; Department of Pediatric Cardiac Center, Anzhen Hospital, Capital Medical University, No.2 Anzhen Street, Chaoyang District, Beijing 100029, China

## Abstract

**Background:** Ambient air pollution is a leading risk factor for cardiovascular diseases. No study has investigated the association between fine particulate matter (PM_2.5_) exposure and prognosis of patients undergoing right ventricle-pulmonary (RV-PA) artery conduit surgery. We hypothesized that PM_2.5_ can lead to stenosis of the conduit by stimulating inflammatory response in the conduit lumen.

**Methods:** From 2013 to 2020, patients with six complicated congenital heart defects and had undergone RV-PA operation in Beijing Fuwai Hospital were selected. Four conduit materials were used: bovine jugular vein graft (BJV), pulmonary homograft (PHG), aortic homograft (AHG), and handmade tri-leaflet expanded polytetrafluoroethylene (ePTFE) conduit. Telephone interviews were used to confirm the patients’ postoperative addresses. The monthly averages of PM_2.5_ concentrations were obtained from the ChinaHighPM_2.5_ dataset using patients’ places of residence. By comparing findings of echocardiography performed prior to patients’ return to their residence and during re-examination, we defined the trans-conduit peak velocity increase of ≥1.5 m/s as the study endpoint.

**Results:** 232 patients were included in the study. Logistic analysis identified the female gender to be a protective factor against conduit velocity increase (Odd Ratio (OR) 0.270 (95% confidential interval (CI): 0.094–0.780), P = 0.016). Compared with BJV conduits, homografts (AHGs and PHGs) (0.052 (95% CI: 0.005–0.558), P = 0.015) and ePTFE conduits (0.009 (95% CI: 0.002–0.054), P <0.001) were protective factors. The cumulative monthly PM_2.5_ concentration (unit 10μg/m^3^) was a risk factor (1.014 (95% CI: 1.001–1.026), P = 0.028). Winter experience was a risk factor (1.971 (95% CI: 1.021–3.804, P = 0.043). In the subgroup analysis, Spearman correlation analysis identified BJV conduits (r = 0.680, P <0.001), PHGs (r = 0.559, P = 0.020), AHGs (r = 0.745, P = 0.021) had medium-to-high positive correlations between the cumulative PM_2.5_ concentration and conduit velocity change. For ePTFE, the correlation was weak and not significant (r = 0.222, P = 0.073).

**Conclusion:** Postoperative PM_2.5_ exposure affects the patency of biologic prosthetic conduits, especially the xenograft. The ePTFE conduit velocity increase is not associated with PM_2.5_ exposure which is a suitable material for patients living in high pollutant concentration areas.

## Introduction

Ambient air pollution has been fully demonstrated to be one of the leading risk factors of cardiovascular diseases. Among the pollutants, fine particulate matter (PM_2.5_), which is <2.5 μm in aerodynamic diameter, is the most harmful [1]. Unfortunately, no studies on PM_2.5_ exposure and postoperative prognosis of patients with congenital heart disease have been conducted. Right ventricle-pulmonary artery conduit connection is a surgical method for reconstructing the right ventricular outflow tract in complex congenital heart defects. Although most patients have good short-term prognosis, the prosthetic conduit may degenerate, mainly due to stenosis, and require replacement or intervention. Previous studies only investigated the clinical factors on conduit durability. The mechanism underlying conduit dysfunction has not been clarified. Since the prosthetic conduit forms a part of circulation after surgery, and PM_2.5_ can reach the circulation through alveoli to cause cardiovascular disease [2], we hypothesize that PM_2.5_ can affect conduit durability. Inhaled PM_2.5_ may stimulate inflammatory response or deposit in the conduit lumen (Fig. 1). Exposure to high concentrations of PM_2.5_ among patients in their postoperative residence may accelerate the conduit stenosis progress. In this study, we identified the effect of patients’ exposure to ambient PM_2.5_ at their postoperative places of residence on conduit patency using data obtained at our institution.

**Figure 1:**
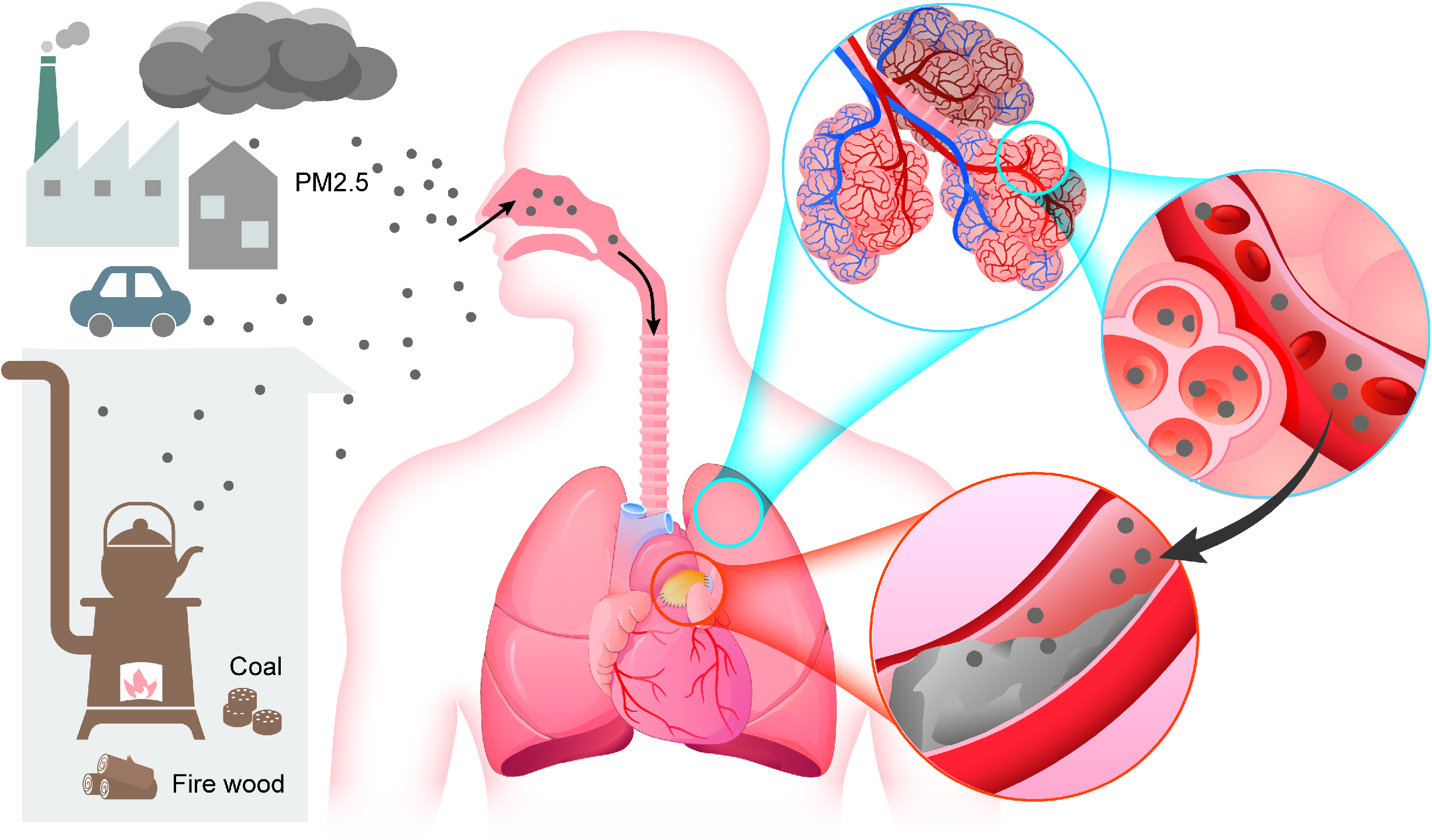
The hypothesis that PM_2.5_ accelerates conduit stenosis. The soluble portion of inhaled PM_2.5_ enters the circulation through alveolar capillaries (blue circle), causing inflammatory response or oxidant stress in the prosthetic conduit, eventually leading to stenosis of the lumen (red circle).

## Methods

### Patients

This retrospective study was conducted in compliance with the Good Clinical Practice (GCP) principle and approved by the Ethics Committee of Fuwai Hospital (ethical number: 2015-651; approved: April 22, 2015). Patients with primary diagnosis of pulmonary atresia (PA), persistent truncus arteriosus (TA), Tetralogy of Fallot (TOF), transposition of the great arteries with pulmonary valve stenosis (TGA+PS), corrected transposition of the great arteries with pulmonary valve stenosis (cTGA+PS), double outlet right ventricle with pulmonary valve stenosis (DORV+PS) and underwent right ventricle-pulmonary artery conduit connection for the first time in Beijing Fuwai Hospital from January 1, 2013 to December 31, 2020 were selected (The Chinese national air pollution monitoring network was established, and data were publicly available from 2013). The need for informed consent from individual patients was waived. During this period, four materials of conduit were used in our institution: bovine jugular vein graft (BJV), pulmonary homograft (PHG), aortic homograft (AHG), and handmade tri-leaflet expanded polytetrafluoroethylene (ePTFE) conduit. Unlike previous studies, this study focused on the association between postoperative PM_2.5_ exposure and the patency of different conduits. Therefore, perioperative factors, such as postoperative acute transplant rejection, inflammatory response, and surgical technique, which may affect conduits, were excluded. Therefore, the exclusion criteria ensured that the selected patients could truly reflect the effect of PM_2.5_ on different material conduits as much as possible. The exclusion criteria were as follows: 1) Patient underwent the single-valved patch procedure; 2) conduit was made of more than one material; 3) death before discharge; 4) conduit already had moderate or severe stenosis (trans-conduit peak velocity ≥3 m/s in echocardiogram results) before the patient returned to place of residence; 5) conduit dilated during follow-up; and 6) conduit velocity increase due to anastomosis site stenosis. Detailed explanations are available in the supplemental materials.

### Endpoint definition

For the same reason, endpoints of previous studies were not applicable for this study. In brief, this study’s endpoint was significant increase in trans-conduit velocity. Two Doppler echocardiography records were compared to define the endpoint. The first record was the trans-conduit peak velocity of the last echocardiogram before the patient returned to residence. The second record was the peak velocity of follow-up examination. If increase in conduit velocity of the two records was ≥1.5 m/s, the increase was defined as significant and reaching the endpoint. The detailed explanations of endpoint design and re-examination choice are available in the supplemental materials.

### Surgical technique

The homografts were harvested and cryopreserved during heart transplantation. Prior to the procedure, patients provided written informed consent. Additionally, the institutional ethics committee approved the procedures. Due to the shortage of homograft donors, conventional BJV grafts have been widely used in our institution and many other institutions in China for the past two decades. However, they were prone to calcification and failure [3]. In the past 5 years, handmade tri-leaflet conduit use has gradually replaced BJV graft use in our institution. The suturing technique for hand-sewn valved conduit was well elaborated in a previous study [4]. Conduit implantations were performed by median sternotomy with cardiopulmonary bypass (CPB). The two ends of the conduit were anastomosed using 6-0 polypropylene sutures to connect the right ventricle and pulmonary artery. The proximal anterior wall of the conduit was prolonged with a pruned autologous or bovine pericardial patch, which was anastomosed with the right ventricular incision to prevent bleeding.

### Monthly average PM_2.5_ concentration measurement

First, we used the coordinate picker system of Baidu Maps to manually convert the actual addresses of each patient obtained through telephone follow-up into latitude and longitude coordinates. These coordinates were then calibrated singly through reverse coordinate comparison. Second, we obtained the monthly PM_2.5_ concentrations across China at a 1-km resolution from the ChinaHighPM_2.5_ dataset [5,6]. The ChinaHighPM_2.5_ dataset is a long-term, comprehensive, high-resolution, and high-quality series of ground-level air pollutant data for China. It is generated from big data (e.g., ground-based measurements, satellite remote sensing products, atmospheric reanalysis, and model simulations) using artificial intelligence by considering the spatiotemporal heterogeneity of air pollution. This dataset is of exceptional quality, with a daily cross-validation coefficient of determination (CV-R2) of 0.92 and a root-mean-square error (RMSE) of 10.76 µgm^-3^. Finally, on the Arcgis platform, we converted the latitudes and longitudes of all involved addresses into point layers and used the “extract value to point” function to extract the monthly ground-level PM_2.5_ concentrations onto the corresponding addresses, thereby obtaining each patient’s monthly PM_2.5_ exposure concentration.

### Data collection and definition

Patients with primary diagnosis of “PA”, “TA”, “TOF”, “TGA+PS”, “cTGA+PS” or “DORV+PS” were identified through our institution’s database. By searching for the word “conduit” in the surgical records, data of patients with these diagnoses that underwent right ventricle-pulmonary artery conduit connection radical surgery for the first time during 2013–2020 were retrieved. By reviewing the surgical records and postoperative clinical records, the patients who might influence the study outcome were excluded. The demographic characteristics and clinical data of the remaining patients were collected. The Z-score calculation in our study was based on the method of Ma et al. [7]. Follow-ups were conducted through telephone interviews. To collect patients’ postoperative information, we designed a questionnaire survey that contained questions on indoor pollution. Details of the questionnaire are available in the supplemental material. Based on follow-up outcomes, patients’ postoperative addresses and change time were determined. Additionally, according to a patient’s location, the monthly average PM_2.5_ concentration at a patient’s residence was measured. The average concentrations for each month of the follow-up period were added to obtain a cumulative value that was used to estimate the PM_2.5_ exposure effect on the patient’s conduit during the whole follow-up period. Since measurements could only be accurate to the monthly average concentration, we had to make a trade-off by using the monthly average concentration of those months during which the patients lived in their places of residence for less than a month. For example, if one returned to one’s residence at the end of the month and lived in the residence for <15 days, then the average concentration for that month was not added to the cumulative value, but to the next month. If the patient stayed for ≥15 days, the average concentration of this month was included. After follow-up, we identified the time when patients returned to their places of residence. A physician reviewed and selected the most appropriate echocardiogram record in clinical data or outpatient data according to the time point.

For the re-examination echocardiogram record, time point selection has been explained in supplemental materials. The physician reviewed and chose the most appropriate re-examination record. For those patients who had no re-examination records in our institution, we requested for images of examinations performed at their local hospitals.

### Statistical analysis

The Kolmogorov-Smirnov method was used for normal distribution test of continuous variables. The variables were presented as mean ± standard deviation, if they were normally distributed. For non-normally distributed data, variables were presented as median and interquartile ranges (IQR). Student’s *t* test or Mann-Whitney U test was used to assess the differences in continuous variables. Categorical variables were presented as frequencies and percentages. The differences were compared using χ^2^ statistics or Fisher’s exact test. Variables with statistical significance in the baseline were selected. After eliminating collinearity variables, the remaining variables were used in a binary logistic regression model for multivariate analysis. All statistical tests were two-tailed, and a P value <0.05 was considered statistically significant. All data were analyzed using SPSS 26.0 (IBM, Chicago, USA). Statistical figures were developed using GraphPad Prism 8.0 (GraphPad, San Diego, USA).

## Results

### Baseline data

From 2013 to 2020, a total of 319 patients with a primary diagnosis of one of the six cardiac anomalies underwent right ventricle pulmonary artery conduit connection for the first time in our institution. After reviewing the surgical records, six patients were identified to have undergone the “single valve procedure”. Five homograft conduits were prolonged with other materials because the length was not enough. Eleven patients died before discharge. Telephone interviews were performed for the remaining 297 patients. Among them, 10 conduits reached moderate or severe stenosis before the patients returned. The conduit velocities of 19 patients increased significantly due to the anastomosis site stenosis. Twenty patients had no re-examination outcomes. Among them, 16 patients died postoperatively. The legal guardians of the four patients who survived did not assent to re-examination. Thirteen patients were lost to follow-up. One patient changed residence frequently, and the parents could not provide the exact times of address change. The diameter of two conduits increased significantly. Finally, 232 patients had accurate postoperative addresses and re-examination records. The flow diagram of patient selection is available in the supplemental material (Fig S1). In all, we implanted BJV, ePTFE, PHG, and AHG conduits in 140, 66, 17, and 9 patients, respectively. Since the AHGs were too few for statistical significance, we combined the AHGs and PHGs into homografts for analysis.

The demographic characteristics, surgical information, and postoperative information are presented in Table 1. In particular, the re-examinations of two patients indicated that the distal portion of the conduits were severely stenosed, leading to the underestimation of the velocity measurement. Additionally, severe conduit stenosis was initially identified in two other patients at their local hospital. Computed Tomography examination in our outpatient department showed severe calcification in the lumen. All these four patients were considered to have reached the study endpoint, and the conduit velocity measured during re-examination was defined that the velocity measured prior to their return to their places of residence adding 1.5 m/s. Finally, 110 patients reached the endpoint during follow-up; among them, 102 (92.7%) patients had BJV conduits. Seventy-six (69.1%) male patients reached the endpoint, compared with 64 (52.5%) male patients in the other group (P = 0.010). The body mass index (BMI) at surgery was greater among patients in the endpoint group (15.99 (14.84, 17.31) vs 15.41 (14.28, 16.77), P = 0.008). Both conduit diameter (18.00 (16.00, 18.00) vs 16.00 (14.00, 16.00), P <0.001) and Z-score (2.06 (1.30, 3.19) vs 1.33 (0.70, 2.16), P <0.001) were greater among patients who did not reach the endpoint. However, in this group, CPB time (175.50 (143.75, 212.00) vs 161.00 (122.75, 189.50), P = 0.001) and crossclamp time (105.23 ± 41.97 vs 94.95 ± 36.39, P = 0.049) were longer. The trans-conduit peak velocity before the patients returned to their places of residence (1.80 (1.47, 2.30) vs 1.50 (1.20, 2.00), P = 0.001) and the velocity during re-examinations (4.20 (3.70, 4.70) vs 2.10 (1.60, 2.53), P < 0.001) were higher in the endpoint group than in the other group. Additionally, the follow-up time was much longer (35.65 (20.73, 55.16) vs 19.25 (11.50, 30.34), P <0.001) in the endpoint group. The cumulative value of all monthly average PM_2.5_ concentrations in the endpoint group was much higher than that in the other group (1688.35 (1120.37, 3191.80) vs 757.25 (411.15, 1193.94), P <0.001). The overall monthly average PM_2.5_ concentration was higher in the endpoint group (51.14 (40.59, 68.68) vs 38.68 (31.11, 49.70), P <0.001). Regarding indoor pollution, more patients in the endpoint group used coal or firewood for winter heating (35 (31.8%) vs 19 (15.6%), P = 0.003) and experienced more winters (3.00 (2.00, 4.50) vs 2.00 (1.00, 2.50), P <0.001).

**Table 1:**
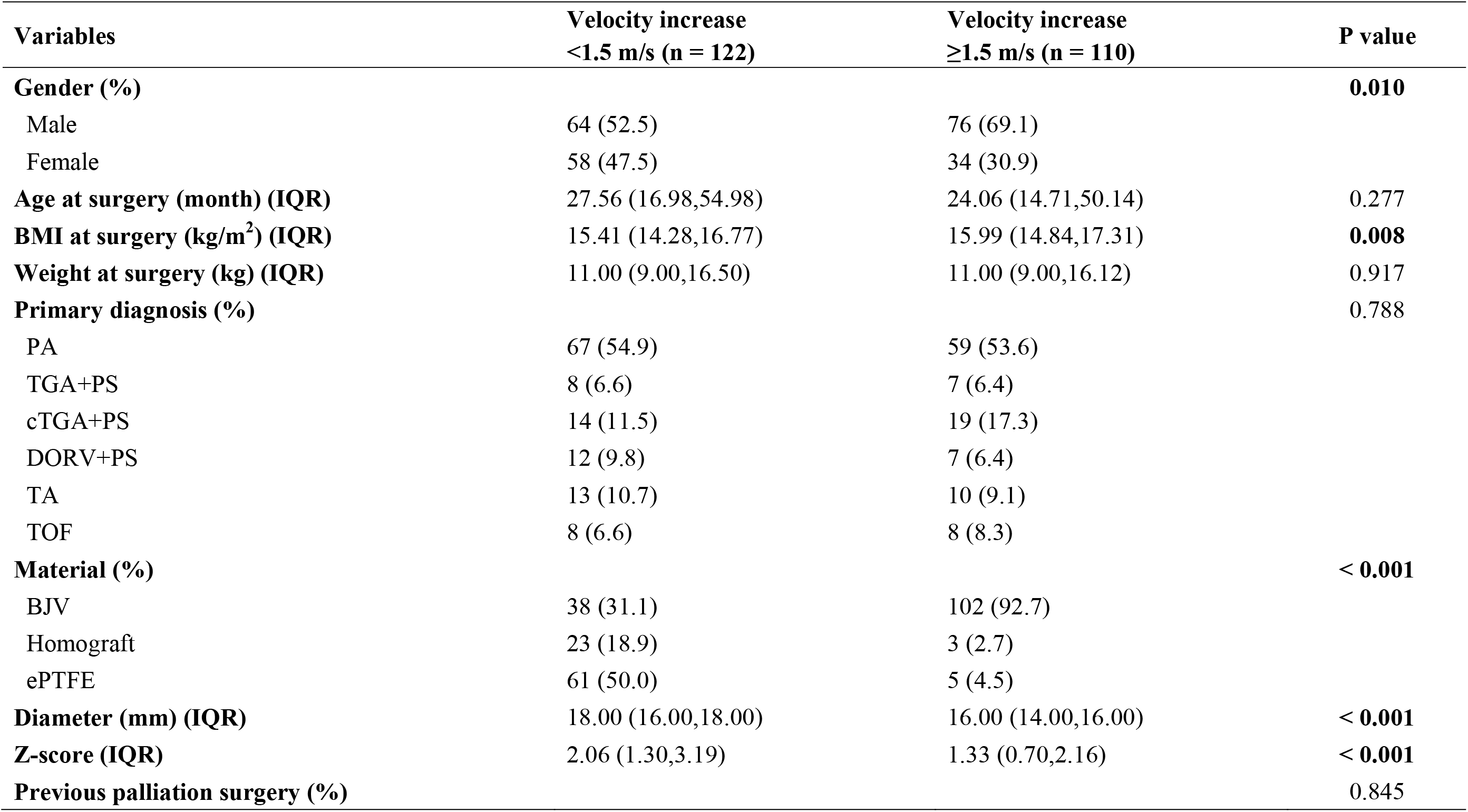
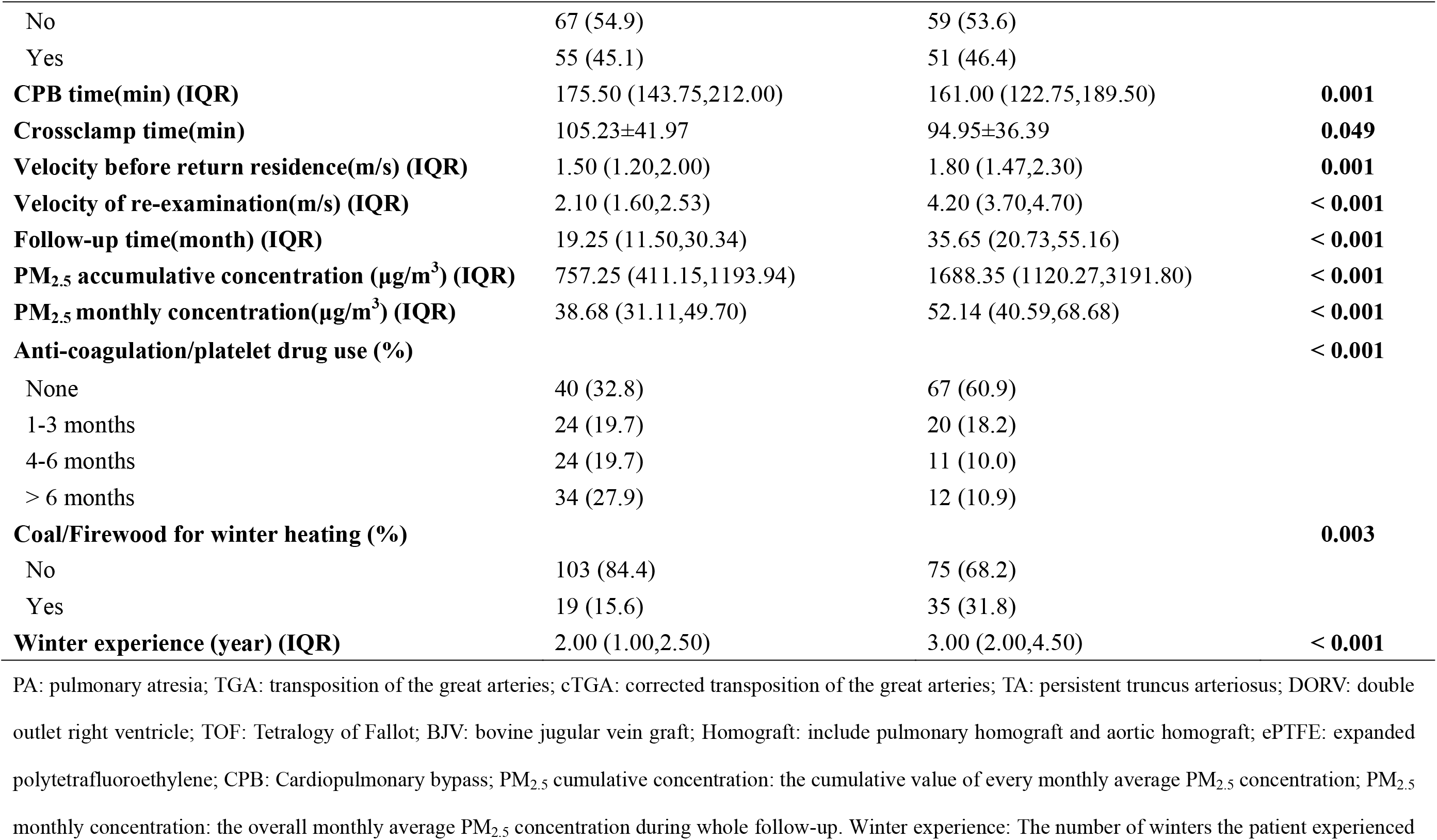
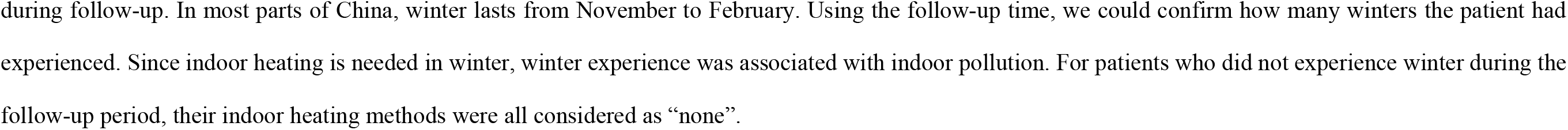
Baseline data.

### Multivariable analysis

The multivariable analysis outcomes are presented in Table 2. A logistic regression model was used to analyze the risk factors for significant increase in conduit velocity. The variables with statistical significance at baseline were selected for analysis. Since velocity before patients returned to their places of residence and velocity during re-examination were variables to determine the study endpoint, they were excluded. Although follow-up time was statistically significant, it had collinearity with cumulative value of PM_2.5_ concentration. Therefore, it was excluded. Finally, multivariate analysis identified female gender to be a protective factor for conduit velocity increase (Odd Ratio (OR): 0.270 (95% confidential interval (CI) 0.094– 0.780), P = 0.016). Regarding conduit material, compared with the BJV conduit, homografts (OR: 0.052 (95% CI: 0.005–0.558), P = 0.015) and ePTFE conduits (OR: 0.009 (95% CI: 0.002 – 0.054), P <0.001) were both protective factors. The cumulative value of monthly average PM_2.5_ concentration was a risk factor for conduit velocity increase. The OR value (1.014 (95% CI: 1.001–1.026), P = 0.028) corresponding to a unit PM_2.5_ concentration was 10 μg/m^3^. Additionally, winter experience, which could be associated with household pollution, was a risk factor (1.971 (95% CI: 1.021–3.804, P = 0.043).

**Table 2:** Multivariable analysis.

| **Variables** | **OR (95%CI)** | **P value** |
| --- | --- | --- |
| **Gender** |  |  |
| Male | REF |  |
| Female | 0.270 (0.094-0.780) | **0.016** |
| **BMI (kg/m<sup>2</sup>)** | 1.060 (0.838-1.340) | 0.628 |
| **Material** |  |  |
| BJV | REF |  |
| Homograft | 0.052 (0.005-0.558) | **0.015** |
| ePTFE | 0.009 (0.002-0.054) | **< 0.001** |
| **Diameter (mm)** | 0.816 (0.616-1.081) | 0.156 |
| **Z-score** | 0.950 (0.583-1.547) | 0.836 |
| **CPB time (min)** | 1.002 (0.986-1.018) | 0.802 |
| **Crossclamp time (min)** | 0.983 (0.963-1.003) | 0.091 |
| **PM<sub>2.5</sub> accumulative concentration (10µg/m<sup>3</sup>) \*** | 1.014 (1.001-1.026) | **0.028** |
| **PM<sub>2.5</sub> monthly concentration (10µg/m<sup>3</sup>) †** | 1.401 (0.940-2.089) | 0.098 |
| **Anti-coagulation/platelet drug use** |  |  |
| None | REF |  |
| 1-3 months | 1.639 (0.427-6.287) | 0.471 |
| 4-6 months | 1.971 (0.424-9.157) | 0.386 |
| > 6 months | 1.280 (0.283-5.792) | 0.749 |
| **Coal/Firewood for winter heating** |  |  |
| No | REF |  |
| Yes | 1.681 (0.515-5.490) | 0.390 |
| **Winter experience (year)** | **1.971 (1.021-3.804)** | **0.043** |

\*†The unit concentration corresponding to the OR value is 10 µg/m<sup>3</sup>; BJV: bovine jugular vein graft; Homograft: includes pulmonary homograft and aortic homograft; ePTFE: expanded polytetrafluoroethylene; CPB: Cardiopulmonary bypass; PM<sub>2.5</sub> cumulative concentration: the cumulative value of every monthly average PM<sub>2.5</sub> concentration; PM<sub>2.5</sub> monthly concentration: the overall monthly average PM<sub>2.5</sub> concentration during the whole follow-up.

### Subgroup analysis

To better reveal the PM_2.5_ effect on different materials of conduits, we further performed subgroup analysis. We combined the AHGs and PHGs into Homografts for baseline data and multivariate analysis. However, in some previous large sample studies, the durability of AHGs was worse than that of PHGs [8–10]. Therefore, we hypothesized that the PM_2.5_ effect on their durability was different. Although the AHGs in our study were few, we still analyzed the two materials.

Scatter plots of cumulative PM_2.5_ concentration and observation months in four type conduits are presented in Figure 2. For BJV conduits, only the cumulative value was low level, the conduit velocity did not increase significantly. For PHGs and ePTFE conduits, most conduit velocities still did not increase significantly when they had exposure to a high cumulative value. For AHGs, one of the nine conduits had the highest cumulative value of PM_2.5_ concentration (850.32 μg/m^3^), and its velocity increased significantly. Therefore, the PM_2.5_ effect on the durability of four material conduits was different. A receiver operating characteristic (ROC) curve was used to obtain the cut-off value of cumulative PM_2.5_ value effect on conduit velocity increase. For BJV conduits, the cut-off value was 916.25μg/m^3^. The area under curve (AUC) was 0.959 (95% CI: 0.929–0.990), P <0.001. The results of other conduits are available in Table S1.

**Figure 2:**
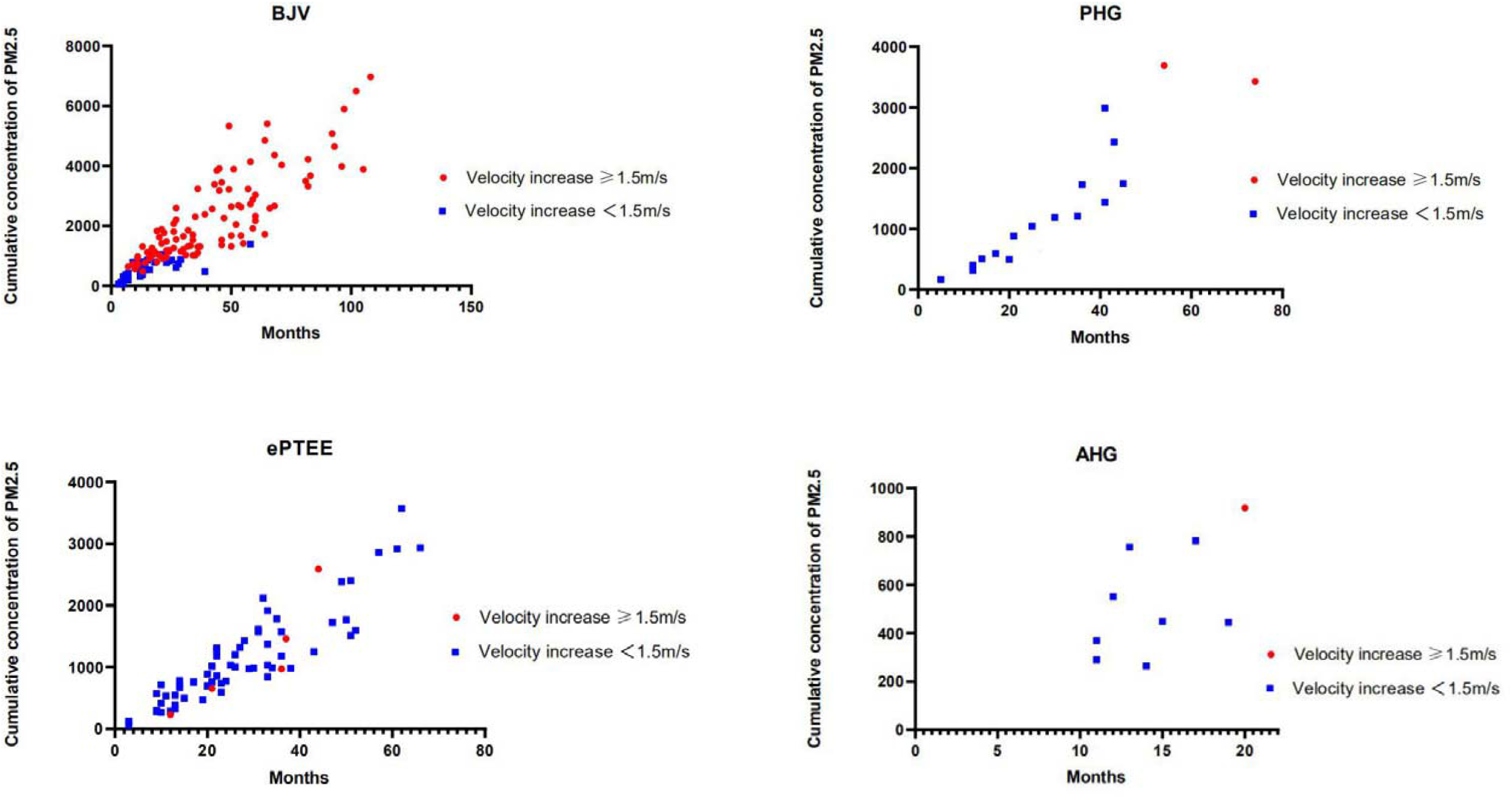
Scatter plot of cumulative PM_2.5_ concentration and observation months for four types of conduit. BJV: bovine jugular vein graft; PHG: pulmonary homograft; ePTFE: expanded polytetrafluoroethylene; AHG: aortic homograft. The unit of PM_2.5_ concentration is μg/m^3^. Since the measurement is accurate to the monthly concentration, the number of months corresponding to each point on the horizontal coordinate is an integer.

For patients in the BJV scatter plot diagram who had high cumulative values and reached the study endpoint, they did not necessarily have high cumulative concentrations before reaching the endpoint. This finding was because in this retrospective study, patients did not undergo re-examinations regularly. Some patients did not undergo re-examinations till they started experiencing symptoms of heart failure. At this point, the conduit velocity acceleration had already become high. For this reason, we analyzed the correlation between cumulative value of PM_2.5_ concentration and conduit velocity increase. Due to the non-normal distribution of the data, Spearman correlation analysis was used to analyze the correlation. The results are presented in Table 3. BJV conduits (r = 0.680, P <0.001), PHGs (r = 0.559, P = 0.020) and AHGs (r = 0.745, P = 0.021) had medium-to-high positive correlations between the two variables. For ePTFE, the correlation was weak and not significant (r = 0.222, P = 0.073).

**Table 3:** Correlation analysis of conduit velocity increase and cumulative value of monthly PM_2.5_ concentration.

| **Material** | **r value** | **P value** |
| --- | --- | --- |
| BJV | **0.680** | **< 0.001** |
| ePTFE | 0.222 | 0.073 |
| PHG | **0.559** | **0.020** |
| AHG | **0.745** | **0.021** |

BJV: bovine jugular vein graft; ePTFE: expanded polytetrafluoroethylene; PHG: pulmonary homograft; AHG: aortic homograft

In summary, the subgroup analysis showed that BJV and AHG were sensitive to PM_2.5_ exposure. The conduit velocity of PHG did not increase significantly until the cumulative PM_2.5_ concentration was at a very high level. Stenosis of ePTFE might not be associated with PM_2.5_ exposure.

## Discussion

### Mechanism of conduit stenosis

Several studies have analyzed the factors influencing the durability of right ventricle – pulmonary artery conduit, but they mostly investigated the perioperative factors, such as material, conduit size (diameter or Z-score), and demographic characteristics of the patient. Although the conclusions of material influence were inconsistent, most studies confirmed that the durability of PHG was significantly longer than those of xenograft and AHG. Besides, many studies found small conduit size, young age, and low-weight of the patient to be risk factors for conduit failure [7,8,10,11]. In our opinion, although these were reasonable risk factors for conduit stenosis, they cannot fully explain the mechanism underlying stenosis progress. During surgery on a young or low-weight patient, surgeons usually choose smaller conduit sizes to match the patient. Compared to the lumen of a large conduit, the limited lumen of a small conduit easily becomes stenosed. The lumen of a large conduit would usually still have more space left during stenosis (Figure 3). Therefore, a larger conduit size prolongs durability. Since conduit diameter should match with the patient’s weight, the chosen conduit size was limited. Only by understanding the mechanism underlying calcification and stenosis would durability of conduit by prolonged.

**Figure 3:**
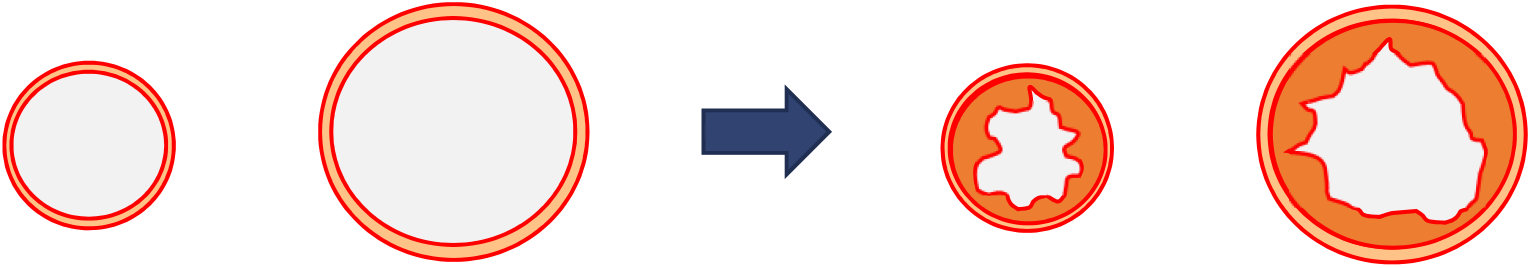
Schematic diagram of the stenosis contrast between large and small conduits Assuming that two different size conduits are exposure to the same degree inflammatory stimulation and become stenosis, the large conduit still has more inner-space than the small one, therefore the conduit velocity change is not obviously.

The American Heart Association published two scientific statements on particulate matter and cardiovascular diseases in 2010 and 2020 [1,2], issuing cautions about the serious harmful effect of PM_2.5_ on various cardiovascular diseases. Inhaled PM_2.5_ can elicit oxidative stress and inflammatory response in lung, systemic circulation, and many other relevant organs [2]. Based on this mechanism, we hypothesized that PM_2.5_ can stimulate inflammatory response to the prosthetic conduit. Chronic pollutant exposure may be a neglected but important risk factor for accelerating the process of stenosis. Furthermore, we suppose this influence should be much obvious in a biologic conduit. Previous studies on pathological analyses of failed xenograft conduits showed chronic inflammatory reactions in stenosed lumen [12]. For ePTFE conduits, pathological changes were proteinaceous infiltration and further calcification of the leaflet but not inflammatory responses [13]. As an inert material, ePTFE exhibits minimal inflammatory and thrombogenic reactions [14]. Due to these characters, we considered its inflammatory response to high concentrations of PM_2.5_ stimulation to be weak. Hence, its durability would be much longer and appropriate for patients living in high pollution areas. Additionally, compared with other materials, BJV conduits, as an implant can cause infective endocarditis (IE), which is a long-term postoperative complication [15]. We believe this may be related to the adhesion of PM_2.5_ to bacteria and toxicants.

According to our data, although the number of homografts used during the time period when PM_2.5_ concentrations were measured at patients’ postoperative places of residence was few, we are still cautious in concluding that PHG is an outstanding material for withstanding PM_2.5_ stimulation. BJV grafts, which were used in our institution, were sensitive to PM_2.5_ exposure. Therefore, BJV is not a recommended choice for right ventricle-pulmonary artery reconstruction. An ePTFE conduit should be used, when no PHG is available as the first choice. However, the mechanism underlying the calcification of ePTFE needs further investigation. According to our data, few ePTFE conduits underwent stenosis early, despite not undergoing any serious pollutant exposure.

### Recommendations for surgeons and patients

In addition to material choice and ambient air pollution, indoor pollution is another risk factor for conduit stenosis, which can be prevented. First, tobacco smoking is one of the main sources of household air pollution that produces a large amount of particulate matter in a short time [16]. Cigarette smoke can be divided into particulate matter and gas-phase; therefore, it has the same mechanism as particulate matter in causing cardiovascular diseases [17]. Compared with ambient particulate matter exposure, the concentration of indoor smoking is not persistently high but much higher in one moment (Figure S2). In addition to smoking, combustion of other substances, such as coal, firewood, and candle can also produce indoor pollutants. In our analysis, the use of coal or firewood for winter heating was not a statistically significant risk factor for conduit velocity increase (OR: 1.681 (95% CI: 0.515–5.490), P = 0.390). This finding was probably due to the simplicity of our estimation of data from the questionnaire; thus, we lack data to quantify the degree of household pollution. From the data obtained from the questionnaires, 216 (93.1%) legal guardians of patients were aware that patients should avoid postoperative exposure to second-hand smoke. However, 54 (23.2%) families still used coal/firewood for winter heating during the follow-up period, and 35 (15.1%) families used coal/firewood for cooking resources. Only seven (3.0%) families had the awareness of using household air-purifiers, although the ambient air quality of their places of residence was serious (overall monthly average concentration median (IQR) was 44.61 (33.06, 50.01) μg/m^3^). On the other hand, to our best knowledge, few studies have investigated the effects of air pollution on patients with congenital heart diseases. Previous studies focused mostly on the association between maternal exposure to ambient air pollution and neonates with congenital heart defects [18]. From observation, both surgeons and patients lack awareness of the harmful effect of air pollution on patients’ prognoses. Protection by patients themselves is even less likely.

### Strengths of the study

This study is the first to investigate and provoke the thought of environmental influence on the conduit of right ventricle-pulmonary artery surgery. The study improved our understanding of why patients with the same/different materials of conduit had different progress of stenosis. Compared with previous studies, this study revealed a neglected but important risk factor for conduit stenosis, providing evidence for surgeons in choosing an appropriate conduit and increasing awareness of patients on the role of household pollution in preventing the progress of stenosis. We strictly screened patients and designed an appropriate study endpoint to truly reflect the effect of PM_2.5_ on conduits. For the measurement of PM_2.5_, we used the most accurate data in China to obtain reliable study outcomes.

### Limitations of study

This retrospective study had some limitations. First, the cumulative value of ambient PM_2.5_ concentration did not represent the exact volume of PM_2.5_ inhaled by patients. The cumulative value was used to coarsely estimate and compare the pollutant exposure of patients in different areas. The exact volume inhaled is concerned with patient’s lung function, activity situation, and other uncertain factors, which cannot be exactly calculated. Second, as mentioned before, the degree of household pollution could not be quantified. Third, similarly, anti-coagulation/platelet drug use could not be quantified. In this retrospective study, patients were administered drugs at the discretion of surgeons but not based on standard therapy. The actual situation was complicated that patients could decrease dosage, withdraw, or change drug. We only measured the duration of drug administration but could not provide detailed data on the category or dosage. Theoretically, drug administration could decelerate conduit degeneration, but the conclusions of current studies are still controversial [19]. Fourth, the sample size of homografts was too small. Further multi-center studies are urgently needed to confirm our findings.

## Conclusions

Postoperative PM_2.5_ exposure affects the patency of biologic conduits used in right ventricle-pulmonary artery surgery, especially the xenograft. The velocity of handmade tri-leaflet ePTFE conduit is an outstanding option, perhaps due to its inert response to pollutants. Both surgeons and patients should be aware of the harmful effect of chronic PM_2.5_ exposure on conduit durability. In the future, further studies should be conducted to enhance the evidence and explore the mechanism of stenosis to improve conduit materials.

## Data Availability

Anyone who wishes to share, reuse, remix, or adapt this material must obtain permission from the corresponding author.

## Acknowledgements

We gratefully acknowledge all the doctors and nurses who participated into the treatment and re-examination of the patients.

## Author contributions

Chao Yue designed the study, collected data, conducted statistical analysis and drafted the manuscript. Jin Li designed the study, measured the PM_2.5_ concentration and drafted the manuscript. Jiaqi Zhang reviewed the echocardiograms. Xu Wang and Qiang Wang contributed to the revision of the manuscript and obtained funding.

